# Forecasting COVID-19 Dynamics and Endpoint in Bangladesh: A Data-driven Approach

**DOI:** 10.1101/2020.06.26.20140905

**Authors:** Al-Ekram Elahee Hridoy, Mohammad Naim, Nazim Uddin Emon, Imrul Hasan Tipo, Safayet Alam, Abdullah Al Mamun, Mohammad Safiqul Islam

**Author notes:** **Correspondence: Dr. Mohammad Shafiqul Islam**, Professor, Department of Pharmacy, Noakhali Science and Technology University.

## Abstract

On December 31, 2019, the World Health Organization (WHO) was informed that atypical pneumonia-like cases have emerged in Wuhan City, Hubei province, China. WHO identified it as a novel coronavirus and declared a global pandemic on March 11^th^, 2020. At the time of writing this, the COVID-19 claimed more than 440 thousand lives worldwide and led to the global economy and social life into an abyss edge in the living memory. As of now, the confirmed cases in Bangladesh have surpassed 100 thousand and more than 1343 deaths putting startling concern on the policymakers and health professionals; thus, prediction models are necessary to forecast a possible number of cases in the future. To shed light on it, in this paper, we presented data-driven estimation methods, the Long Short-Term Memory (LSTM) networks, and Logistic Curve methods to predict the possible number of COVID-19 cases in Bangladesh for the upcoming months. The results using Logistic Curve suggests that Bangladesh has passed the inflection point on around 28-30 May 2020, a plausible end date to be on the 2^nd^ of January 2021 and it is expected that the total number of infected people to be between 187 thousand to 193 thousand with the assumption that stringent policies are in place. The logistic curve also suggested that Bangladesh would reach peak COVID-19 cases at the end of August with more than 185 thousand total confirmed cases, and around 6000 thousand daily new cases may observe. Our findings recommend that the containment strategies should immediately implement to reduce transmission and epidemic rate of COVID-19 in upcoming days.

**Highlights:**

1. According to the Logistic curve fitting analysis, the inflection point of the COVID-19 pandemic has recently passed, which was approximately between May 28, 2020, to May 30, 2020.
2. It is estimated that the total number of confirmed cases will be around 187-193 thousand at the end of the epidemic. We expect that the actual number will most likely to in between these two values, under the assumption that the current transmission is stable and improved stringent policies will be in place to contain the spread of COVID-19.
3. The estimated total death toll will be around 3600-4000 at the end of the epidemic.
4. The epidemic of COVID-19 in Bangladesh will be mostly under control by the 2nd of January 2021 if stringent measures are taken immediately.

## Introduction

Coronavirus disease 2019 (COVID-19), the ongoing worldwide pandemic, is a highly contagious disease that predominantly causes respiratory complications ranging from the cough to acute respiratory distress syndrome. At the time of writing this, it has claimed more than 4,46000 lives, spread in 213 countries around the world. Since the 8^th^ December 2019, atypical pneumonia cases have identified in Wuhan City, Hubei province, China (Zhu et al., 2020), later this disease broke out within China and continued to spread all over the world, causing startling panic worldwide. Since then, it poses unprecedented challenges to global health and a challenging task for the scientific community to identify its behavior. In the following months, the World Health Organization (WHO) declared it a global pandemic due to its highly contagious behavior (Wang, Wang, Ye, & Liu, 2020). The virus severe acute respiratory syndrome coronavirus-2 (SARS-CoV-2), named by the International Committee on Taxonomy of Viruses (ICTV), that causes COVID-19 belongs to genus Beta coronavirus, and it is a positive-sense single-stranded RNA (+ssRNA) virus and thus can mutate frequently to cope up with changing environments (Kaneko, Nimmerjahn, & Ravetch, 2006). The mean incubation period of COVID-19 is estimated 6.5 days (2-14 days) while patients remain asymptomatic or experience a little symptom and thus spreading the virus silently (Lai, Shih, Ko, Tang, & Hsueh, 2020; Rothan & Byrareddy, 2020). As of now, there is no proven vaccine of COVID-19, that altogether contributed to its seriousness. Symptoms of COVID-19 vary from person to person. However, common symptoms include respiratory stress, fever, cough, shortness of breath, and breathing difficulties (Yang et al., 2020). It is estimated that the case-fatality rate of COVID-19 is 2.3%, but older people with comorbidities, the rate is higher (Novel, 2020). WHO (2020) recommended that preventive measures for COVID-19 includes maintaining social distancing, washing hands frequently, avoiding touching the mouth, nose, and face. The first three cases of COVID-19 was reported in Bangladesh on 8^th^ March 2020 origin from Italy. It spreads to the maximum of the districts of the country within a month. After the confirmation of the first three cases of COVID-19 on 8^th^ March 2020, the government of Bangladesh declared a 10 days lockdown from 26^th^ March 2020 to 4^th^ April 2020. However, lockdown periods were later extended several times until 30^th^ May 2020, and then zone-based lockdown strategies have been taken. In addition, All the educational institutions closed down from 17^th^ March 2020 and still ongoing. The government of Bangladesh had suspended all national and international flights from 30^th^ March 2020 and reopened international flights on a small scale from 16th June 2020.

At the time of writing this, the confirmed cases in Bangladesh have surpassed 100 thousand and more than 1343 death toll. The administration is struggling to accommodate COVID-19 patients. Therefore, some predictive models will be an excellent tool to predict future cases in upcoming days (Tobías, 2020; L. Wang et al., 2020). The COVID-19 data depends on non-linear function; thus, using linear methods might fail to capture the COVID-19 dynamics (Chimmula & Zhang, 2020). Several statistical methods such as Auto-Regressive Moving Average (ARIMA), Moving Average (MA), Auto-Regressive (AR) methods have been used to understand the COVID-19 dynamics (Benvenuto, Giovanetti, Vassallo, Angeletti, & Ciccozzi, 2020; Dehesh, Mardani-Fard, & Dehesh, 2020) but these methods did not fit well COVID-19. To overcome such barriers, we adopted data-driven Deep Learning-based LSTM networks and Logistic Curve fitting methods to predict future likely cases and deaths. The proposed methods approximately fit real data; thus, policymakers can prepare to take the rush of COVID-19 in upcoming days. Besides, early prediction using mathematical models would help government officials of Bangladesh and also the health administration of Bangladesh to prepare beforehand and take the precautionary measures to compensate for the causalities of COVID-19 Pandemic. Mathematical models are often used to understand infectious disease dynamics. Several initial studies were performed for COVID-19 transmission. Reproduction Number (*R*_0_) is often used in order to model infectious disease. An *R*_0_ is, basically, how many people can get the infection by the infected person. An *R*_0_ value 3 represent infected people will infect 3 persons around him/her. Liu, Gayle, Wilder-Smith, and Rocklöv (2020) estimated initial *R*_0_ of COVID-19 in China was 4.2. The *R*_0_ for the Diamond Princess, a cruise ship was estimated at 2.1 (Zhao et al., 2020). However, such an approach often leads to difficulties in predicting future cases. The Logistic Growth model and the Susceptible-Infectious-Recovered (SIR) model often use for epidemiological prediction and also have been employed on COVID-19 data. Roosa et al. (2020) used a Generalized Logistic Growth model to assess short-term forecasts of the total confirmed cases in China. Vattay (2020), using the Logistic Growth model, predicted the ultimate death numbers in Hubei, China and Italy. Zhou et al. (2020), using the Susceptible-Exposed-Infectious-Recovered (SEIR) and Logistic Growth model, forecasted the epidemic size of eight different countries. Rahman, Ahmed, Hossain, Haque, and Hossain (2020) using SIR models projected final infection size in Bangladesh in assuming eight different scenarios. al Azad and Hussain (2020) using the SIR model estimated that the *R*_0_ of COVID-19 in Bangladesh as of May is 2.88.

However, reviewing the literature, it is observed that a data-driven approach to the prediction of COVID-19 dynamics is limited in Bangladesh. Therefore, this study adopted a deep learning method LSTM and Logistic Curve model to predict likely future cases of COVID-19, its peak time, and the plausible end-date of the COVID-19 pandemic in Bangladesh. Our analysis may help policymakers and health professionals to take necessary measures toward COVID-19 in upcoming months.

The layout of this article is as follows: In section 2, similar studies have been presented. In section 3, methods and models have been explained. In section 4, results and discussion has been discussed, and in section 5, concluding remarks have been presented.

## Related Works

Several works had been done globally with Machine learning approaches since the beginning of the pandemic COVID-19, some of them are mentioned below:

Chimmula and Zhang (2020), using the LSTM model, they predicted the possible ending point of the Pandemic COVID-19 outbreak in Canada would be around June 2020.

Kafieh et al. (2020) used a different machine learning approach to predict the future number of cases in Iran and found that M-LSTM was the most accurate model for their study.

Tomar and Gupta (2020) used the LSTM and curve-fitting model for prediction of the number of COVID-19 cases in India 30 days ahead and showed the effect of preventive measures like social isolation and lockdown on the spread of COVID-19.

Hu, Ge, Jin, and Xiong (2020) developed Modified Auto-Encoders (MAE) to forecast the number of accumulative and new confirmed cases of Covid-19 in China.

Fountain-Jones et al. (2019) used Machine learning models to predict disease risk in animal populations by pathogen exposure.

## Methods & Models

### Dataset

The COVID-19 data used in this research is collected from Johns Hopkins University’s GitHub repository (https://github.com/CSSEGISandData/COVID-19). The dataset also includes the number of total confirmed cases, fatalities, and recovered patients by the end of each day. This study has been used the COVID-19 data of Bangladesh from 8^th^ March 2020 to 13^th^ June 2020 for LSTM models. The deep learning models are trained with 98 occurrences of total confirmed, recovered and death COVID-19 cases. The dataset is divided into 97% training set and 3% test set. For the Logistic Curve fitting model, data from 8^th^ March 2020 to 18^th^ June 2020 has been used.

### Time series (TS)

A sequence of data points over a regular interval of time is called time-series (TS). In the regression predictive modeling approach, it is desired that the observation is in temporal structure. In other words, it will remain consistent over time. In TS terminology, the consistency is referred to as time series being stationary, which indicates it has constant mean and variance in respect to time. These are important characteristics in TS; however, it can be easily violated by presences of trend, seasonality, and error in TS. A TS is said to have a trend when a certain pattern repeats over regular intervals of time. A non-stationary time series has trends, seasonal effects; as a result, its statistical properties change over time. Time series datasets of COVID-19 may have non-stationary patterns due to lockdown, social distancing, etc. Thus, it is important to know the nature of TS before applying forecasting methods on the given TS dataset. This study adopted the Augmented Dickey-Fuller (ADF) test (Cheung & Lai, 1995) to check the nature of TS. ADF test checks the stationarity of TS dataset. ADF is a unit root test that checks the impacts of trends in a given TS. The results are interpreted by the p-value from the test. If the p-value is between a threshold of (1-5) %, it suggests that we can reject the null hypothesis (i.e., it does not have a unit root, and it can be regarded as stationary series); otherwise, a p-value above the threshold suggests we fail to reject the null hypothesis (i.e., it does have a unit root, and it can be regarded as non-stationary series). The ADF test of our study suggests we fail to reject the null hypothesis; the data has a unit root, and data is non-stationary.

### Long Short-Term Memory (LSTM) Network

Recurrent neural network (RNN) is a type of artificial neural network, which is used in temporal domains to learn sequential patterns. Recurrent LSTM networks can address the limitations of traditional time series forecasting techniques by adapting nonlinearities of given COVID-19 dataset and can result in a state of the art results on temporal data (Chimmula & Zhang, 2020). The Long Short-Term Memory model (LSTM) (M. Zhang, Geng, & Chen, 2020; Q. Zhang, Gao, Liu, & Zheng, 2020) is an advancement from the recurrent neural network. However, RNN suffers from vanishing gradient problems, meaning networks cannot learn from long data sequences. To overcome this barrier, the LSTM was proposed by (Hochreiter & Schmidhuber, 1997). Each block of LSTM operates at different time steps and passes its output to the next block until the final LSTM block generates the sequential output (**Figure 2)**. The basic structure of LSTM consists of four gates-input gate, forget gate, control gate and output gate.

**Figure 1.**
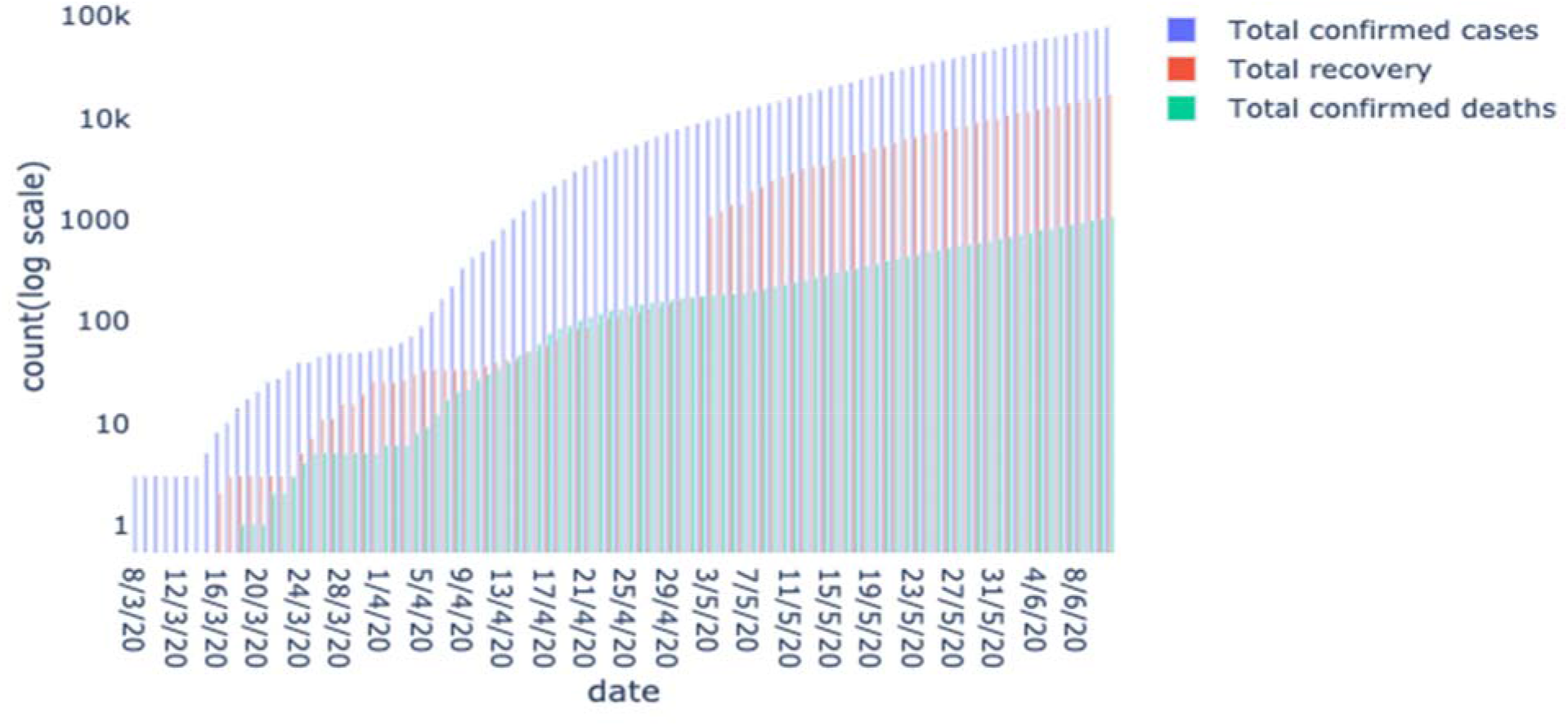
Total confirmed, recovered and death cases in Bangladesh up to June 8, 2020

**Figure 2.**
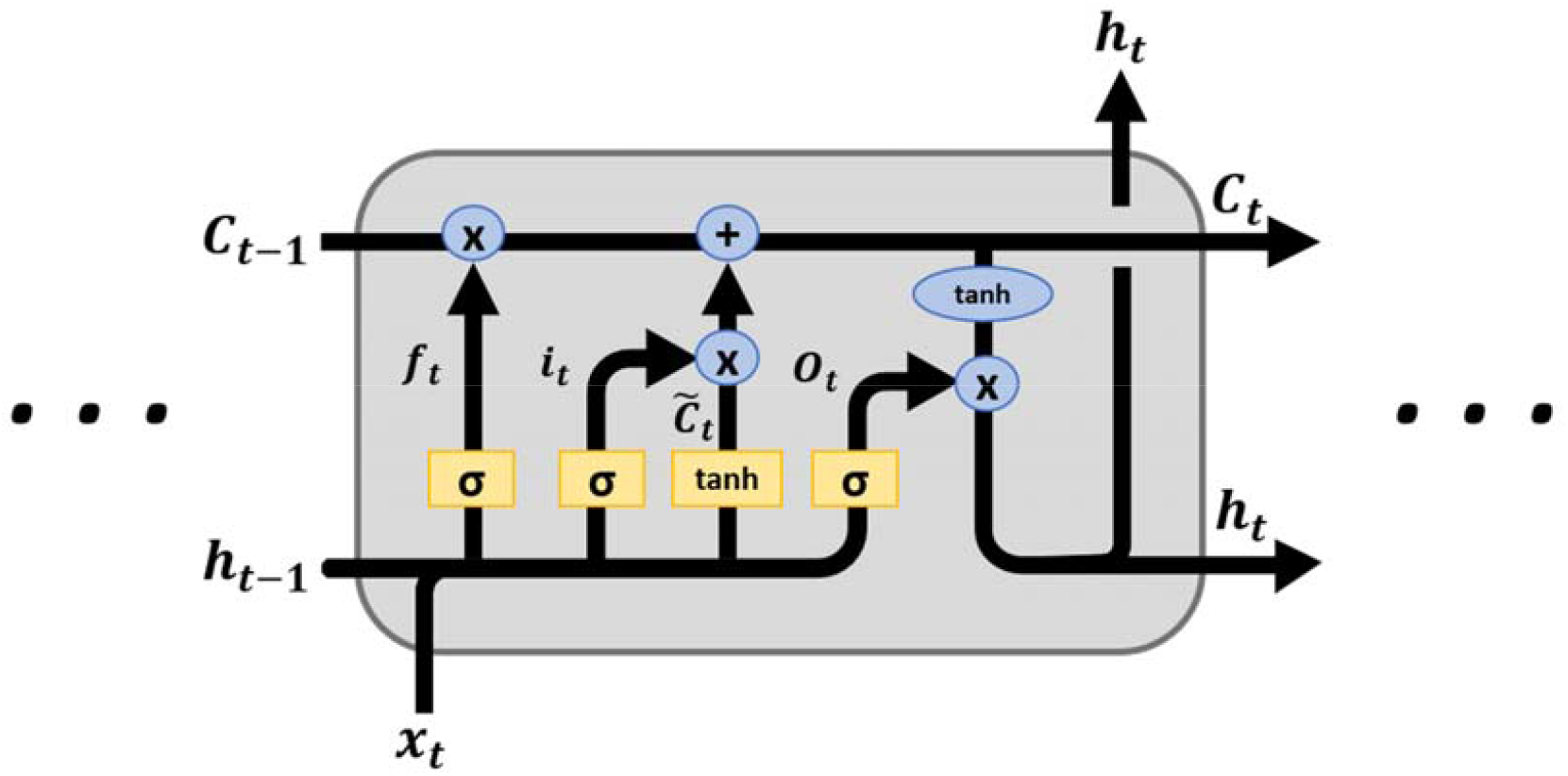
basic LSTM cell structure

In figure 2, *i*_*t*_, *f*_*t*_, *C*_*t*_, *O*_*t*_ are the input gate, forget gate, control gate and output gate, respectively. The details of these four gates are enlightened below:

The input gate decides which information can be transferred from the previous cell to the current cell. The input gate is defined as

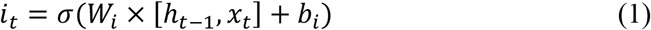

The forget gate decides it should store the information from the input of previous memory or not. The forget gate is defined as

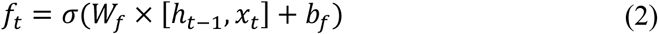

The control gate controls the update of the cell and is defined as

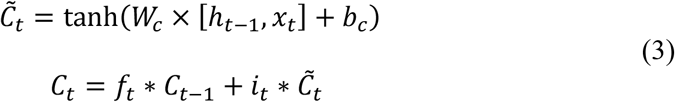

Finally, the output gate updates the hidden layer (*h*_*t*-1_) and also updates the output and is given by the following equations:

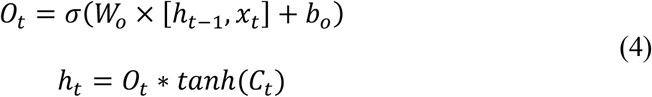

In the above equations, *x*_*t*_ is the input and *W and b* are the corresponding weight matrices and bias, respectively; *C*_*t*-1_ and *C*_*t*_ are the previous and current block memory, respectively; *h*_*t*-1_ and *h*_*t*_ are previous and current blocks the output, respectively. Moreover, *tanh* is the hyperbolic tangent function that used to scale the values into range -1 to 1 and *σ* is the sigmoid activation function, which gives the output in between 0 to 1.

### Logistic Curve Fitting Model

The Logistic Curve model is widely used to describe the growth of a population. An infectious disease outbreak like COVID-19 can be seen as the growth of the population of a pathogen agent; thus, a logistic model seems reasonable to model the progression of the agent (Bertolaccini & Spaggiari, 2020). The most generic expression of a logistic function is given below:

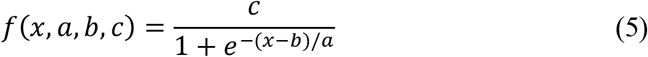

Where *x* is the time and *a, b, c* are parameters; *a* refers to the infection speed, *b* is the day with the maximum infections occurred and *c* is the total number of recorded infected people at the infection’s end.

### Evaluation Metrics

Evaluating the model accuracy is a crucial part of a machine learning model, which gives us the idea of how well the model is predicting. There are many types of evaluation matrices and vary according to the model type. It is wise to evaluate a model with multiple metrics to ensure the correct and optimal operation of the model. In our study, the following evaluation matrices have been considered:

### Mean Squared Error (MSE)

MSE metric represents the difference between the true and predicted values extracted by squared the average difference over the dataset. The equation of MSE is given below:

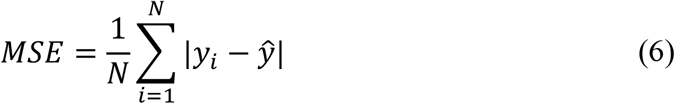

### Root Mean Squared Error (RMSE)

RMSE is nothing but the error rate by the square root of MSE and is defined by

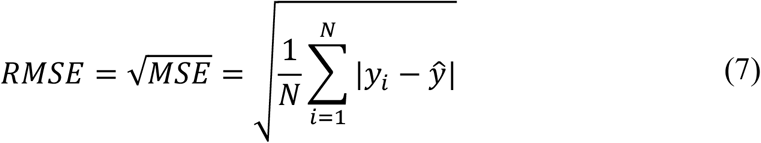

### R-squared (*R*^2^)

The *R*^2^ or the coefficient of determination metric represents the coefficient of how well the predicted values fit compared to the true values of the dataset. The value from 0 to 1 interpreted as percentages where higher value means the better model. The equation is given below:

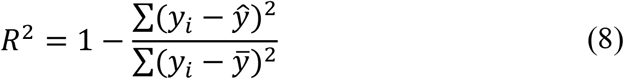

In the above equations, *y*_*i*_ is *i*^*th*^ indexed value, *ŷ* and 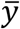 are the predicted value and the mean value of *y*, respectively.

### Mean Absolute Error Percentage (MAPE)

MAPE metric is used to measure the size of the error in percentage terms regarding the actual values and is defined as

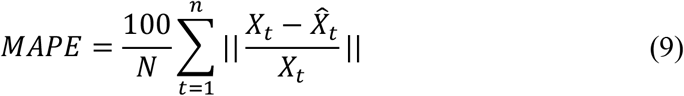

where *X*_*t*_ is the actual value and 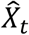 is the corresponding estimated value for *t*^th^ sample from all n available samples.

## Results and Discussion

In the following section, we present the experimental results for the validation and analysis of the proposed model introduced in this study. We hope that our approaches and predictive models will shed some light on the COVID-19 dynamics in Bangladesh and may help us to be aware of upcoming undesired circumstances and take necessary measures in advance and possible suppression of the COVID-19 Pandemic in Bangladesh.

### Data-driven approach

This study has been used the COVID-19 data of Bangladesh from 8^th^ March 2020 (When the first case of COVID-19 was registered in Bangladesh) to 13^th^ June 2020. The deep learning models are trained with 98 occurrences of total confirmed, recovered and death COVID-19 cases. The dataset is divided into 97% training set (from 8^th^ March 2020 to 10^th^ June 2020) and 3% test set (from 11^th^ June 2020 to 13^th^ June 2020). For Logistic Curve fitting data from 8^th^ March 2020 to 18^th^ June 2020 has been considered. The performance evaluation of LSTM models is examined with mean square error (MSE), root mean square error (RMSE) and mean absolute percentage error (MAPE). Before selecting the best-performed LSTM model, we considered three types of LSTM models (i.e. Vanilla, Stacked, Bidirectional) for univariate time series forecasting. We executed each model for total confirmed, recovered and deceased data, then the best-performed model is considered based on performance metrics. The performance results of different LSTM models are described in Table 1.

**Table 1.** performance results of different LSTM models

| **Total Confirmed** |  |  |  |  |
| --- | --- | --- | --- | --- |
| **LSTM Model(s)** | **MSE** | **RMSE** | **$R^2$** | **MAPE** |
| Vanilla | 3164020 | 1778.77 |  |  |
| **Stacked** | **352556.652** | **593.764** | **0.95** | **1.76%** |
| Bidirectional | 4040000 | 2009.970 |  |  |
| **Total Recovered** |  |  |  |  |
| Vanilla | 842289 | 917.763 |  |  |
| Stacked | 1240980 | 1113.990 |  |  |
| **Bidirectional** | **54610.71** | **233.689** | **0.72** | **1.18%** |
| **Total Death** |  |  |  |  |
| Vanilla | 5499.64 | 74.1595 |  |  |
| Stacked | 1359.87 | 36.876 |  |  |
| **Bidirectional** | **406.60** | **20.164** | **0.75** | **1.25%** |

From Table 1, we can observe that for total confirmed cases, the stacked LSTM model performed better than the vanilla and bidirectional LSTM model. The RMSE score of selected stacked LSTM model for total confirmed cases is 593.36, *R*^2^ score 0.95 and accuracy 98.24%. For total recovered, the bidirectional LSTM model performed better than others; the RMSE is 233.68 and *R*^2^ score 0.72 and accuracy 98.82%. For total death, bidirectional LSTM performed better than others; the RMSE is 20.16, *R*^2^ score 0.75, and accuracy 98.75%.

LSTM results suggest that in the near future, the total numbers of confirmed cases will continue to grow exponentially. Thus, strict-lockdown and maintaining social distancing is necessary to reduce the transmission of COVID-19. The predicted data from LSTM model is very close to actual.

### Logistic Curve fitting Model

Traditional infectious disease modeling uses differential equations based on the dynamics of the diseases where too many factors are also associated; thus, it often leads to over-fitting. It is observed that the Logistic Curve fits the COVID-19 data of Bangladesh very well. The parameters of the Logistic Curve are obtained by using a non-linear optimization algorithm, and results are shown in Table 2. The COVID-19 data of Bangladesh from 8^th^ March 2020 to 18^th^ June 2020 has been considered for Logistic Curve fitting.

**Table 2.** parameters obtained for total confirmed cases using the logistic model with standard errors.

| Parameter | $a$ | $b$ | $c$ | $R^2$ | Plausible end of the infection |
| --- | --- | --- | --- | --- | --- |
| Total confirmed cases | 14.702 ± 0.187 | 15/06/2020 | 187169 ± 5610 | 0.99918 | ∼ 02/01/2021 |
| Total deceased cases | 18.262 ± 0.365 | 16/06/2020 | 3692 ± 311 | 0.99810 | |

### Total number of confirmed cases

As of 18^th^ June 2020, the total confirmed cases of COVID-19 in Bangladesh are 102,292 and ranked in 17^th^ most infected countries in the world. The number of recorded cases has increased dramatically between 40-50 days since the first confirmed cases, which represents the sudden changes from where the number of infected cases started following an exponential trend. It is observed that the Logistic curve fitting model (LG) approximates the total number of confirmed cases very well. The fitting parameters are included in Table 2, where a is the infection speed, b is the day with maximum infections occurred and c refers to the total number of confirmed infected people at the infection’s end and *R*^2^ is the fitting goodness of the total confirmed cases.

The logistic curve of Figure 4 suggests that Bangladesh is approaching its peak. Since the data is approximately fitting with the curve, this study suggests that assuming the people will maintain social distancing and the government of Bangladesh will keep various kinds of interventions, at the end of June 2020, there will be 135-140 thousand total confirmed cases. At the end of July 2020, there will be 178-180 thousand total confirmed cases, and at the end of August 2020, there will be more than 186 thousand confirmed cases, where the attainment of peak cases will be reached. Figure 4 also suggests that the curve will flatten at the beginning of September 2020 if strict-lockdown in place. The plausible end of the infection is at the beginning of January 2021 with total confirmed cases in between 187 to 193 thousand.

**Figure 3.**
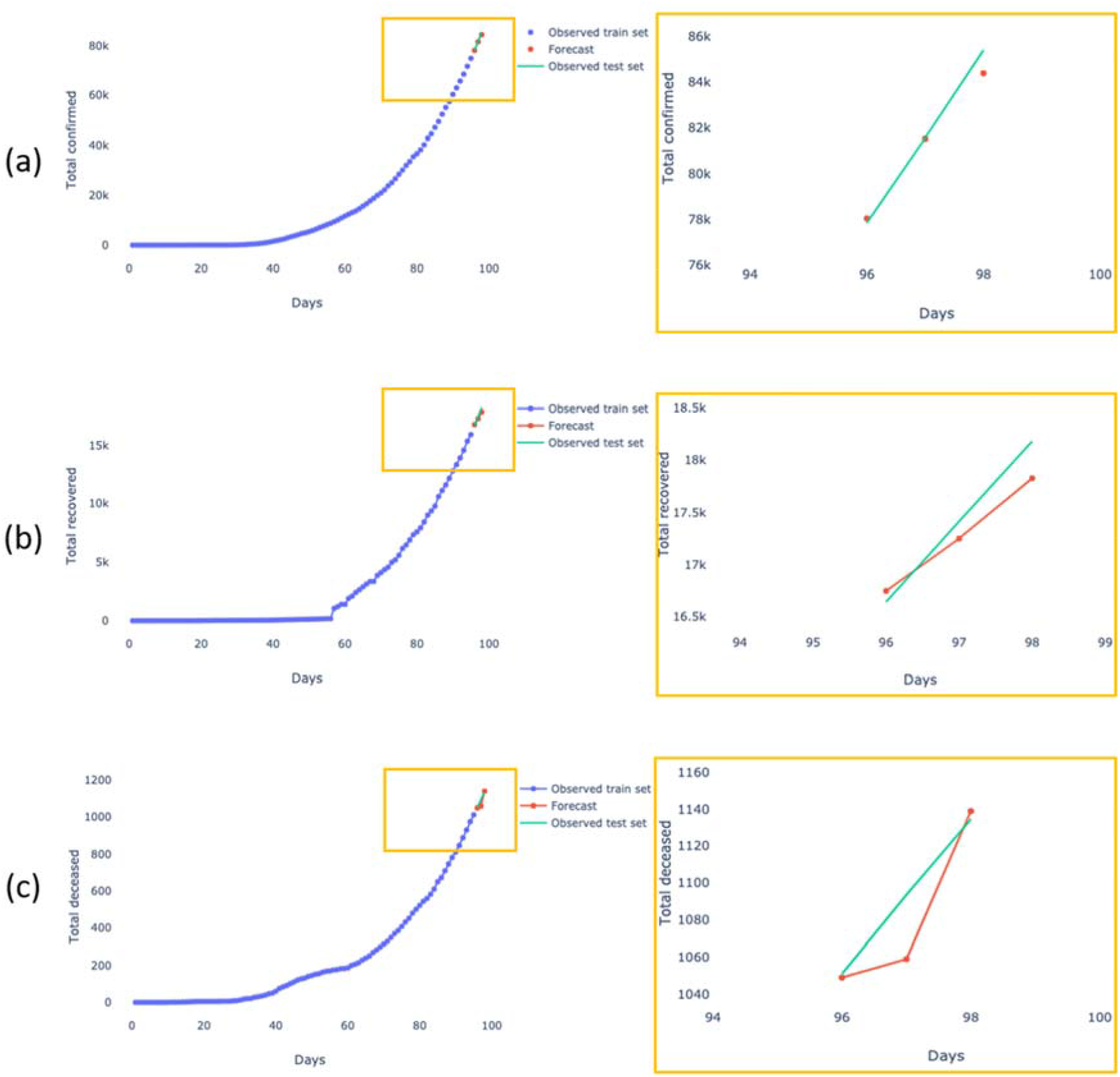
(a) Total number of confirmed cases prediction using LSTM (b) Total number of recovered cases prediction using LSTM (c) Total number of deceased cases prediction using LSTM.

**Figure 4.**
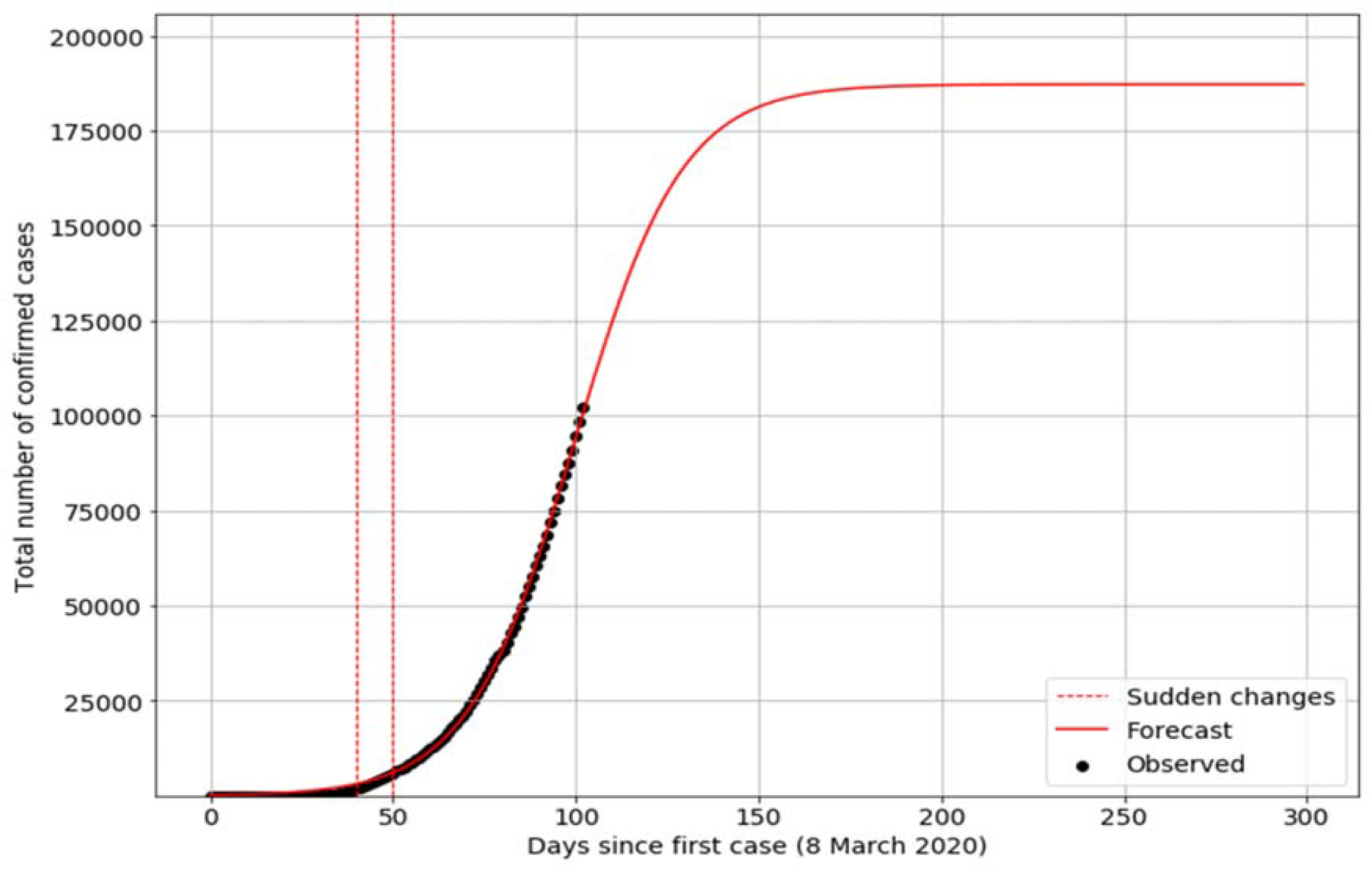
Logistic Curve fitting on the total number of confirmed cases

### Inflection point

Turning point or inflection point is the point where the exponential growth of transmission rate starts to decrease; more formally, its concavity changes from upwards to downwards. It is the midpoint of the spread of an infectious disease. In order to know the inflection point, we used Logistic curve-fitting on the new confirmed cases in Figure 5. Based on Figure 5, Bangladesh has recently passed the rising inflection point and approaching the peak of the infections.

**Figure 5.**
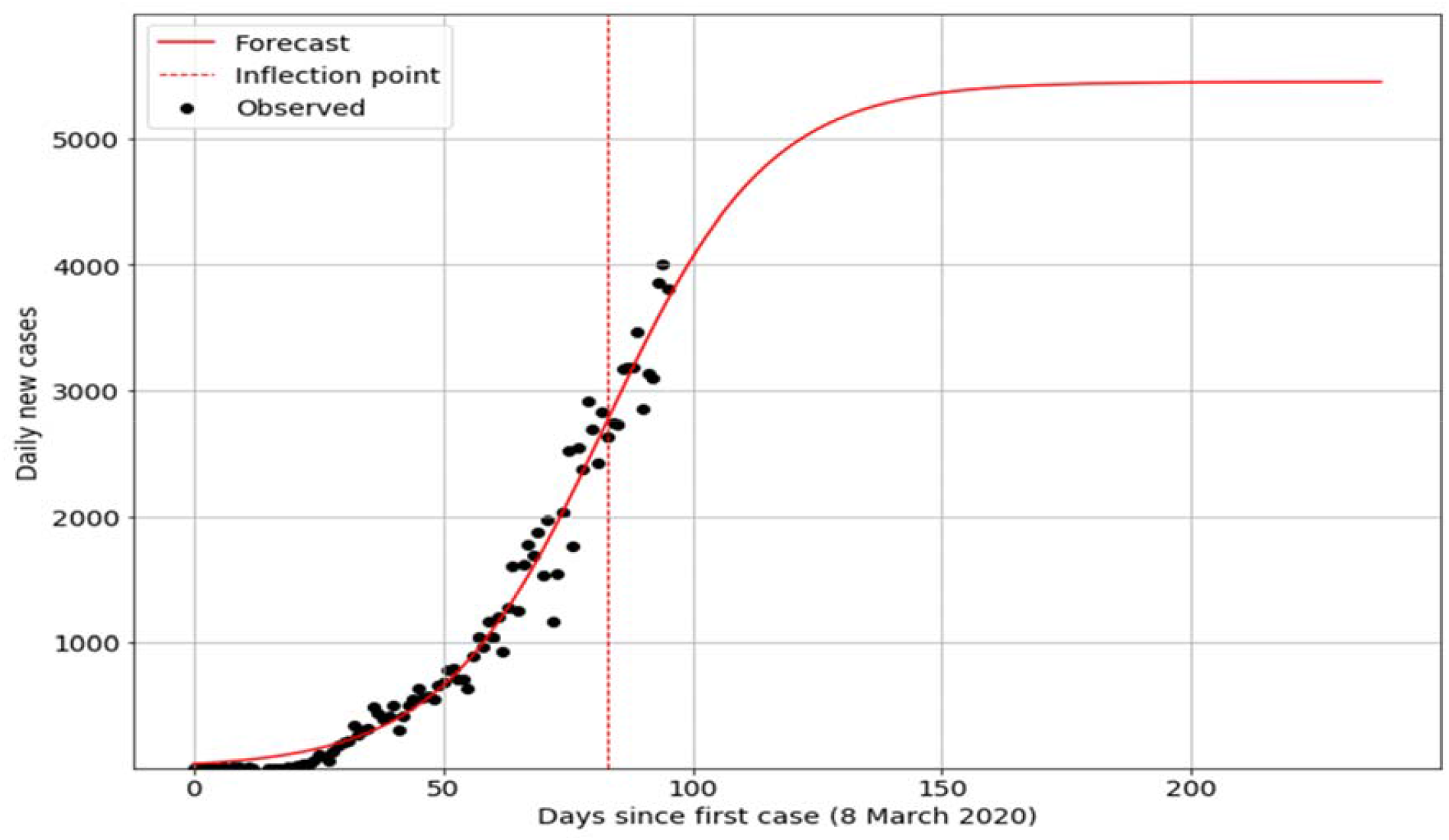
Logistic Curve fitting on the total number of new cases

**Figure 6.**
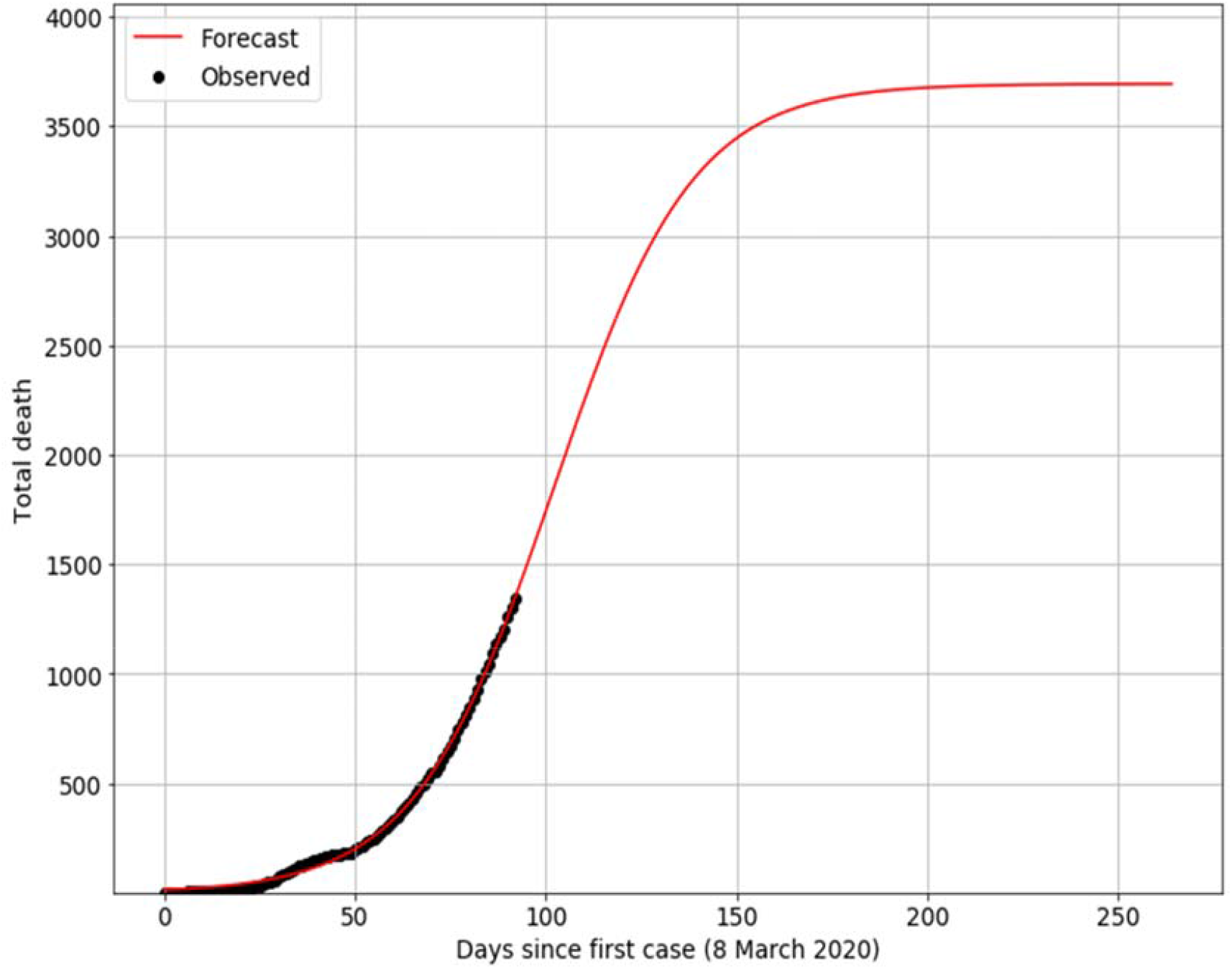
Logistic Curve fitting on the total number of death cases.

Figure 5 also suggests that a rising inflection point was passed around May 28 – 30, 2020. It was the 83-85 days later since the first confirmed cases. Since Bangladesh has recently passed the inflection point in terms of the COVID-19, it indicates that COVID-19 cases are nearing its peak. Based on Figure 4 and 5, the Logistic Curve predicted that the highest peak will be around at the end of August 2020 with more than 186 thousand confirmed cases, where daily new cases may surpass more than 6,000. However, it is worth noting that transmission of the COVID-19 depends on several exogenous factors such as lockdown, social-distancing; thus, this prediction might slightly vary to reality.

### Total death toll

Logistic model used to predict the total death toll in Bangladesh in upcoming days.

It is observed that the Logistic model approximates the total number of death tolls very well. The fitting parameters are included in Table 2, where a is the death speed, b is the day with maximum deaths occurred, and c refers to the total number of deaths at the infection’s end and *R*^2^ is the fitting goodness of the total deaths. The LG curve suggests that at the end of June 2020, there will be around 200 death tolls, and at the end of July 2020, there will be more than 3150 death tolls. The maximum death toll will reach around the end of August 2020, with more than 3600 total death cases. Since the number of total deaths depends on the health care capacity of a country, we suspect this number will vary to reality.

## Conclusion

In this study, a data-driven forecasting/estimation method has been adopted to estimate possible total confirmed and deceased cases in upcoming months. The rate of transmission in Bangladesh, still, is following exponential growth despite taking several intervention strategies by the government of Bangladesh. This is the first study, to best of our knowledge, to model COVID-19 transmission in Bangladesh using a deep learning approach. However, it worth noting that despite of having small-dataset, our model gave promising accuracy. Thus, these models will help policymakers to understand the future of the COVID-19 trajectory in Bangladesh and implementing interventions as well as estimate the impact of interventions. Our LSTM models can give valuable insights into the COVID-19 transmission in Bangladesh and possibly predict future outbreaks. In addition, we also adopted the Logistic curve fitting model to estimate confirmed cases and the death toll. However, it worth noting that the prediction and conclusion drawn from the results are under the preconditions that control measures for the COVID-19 in Bangladesh are stable and reliable, and the virus will not mutate as well in the future. The conclusion has drawn from fitting 103 days data where 65 days (25^th^ March 2020 to 30^th^ June 2020) of nation-wide lockdown was in place. The prediction will improve if new data are generated in the upcoming days.

## Data Availability

All data were included in this study and on request, further information will be given by the corresponding author.

## Acknowledgment

We like to thanks all the front-line fighters-doctors, pharmacists, biochemists, microbiologists, nurses, medical workers, and police, who are putting their lives at risk to keep us safe.

## Ethical Consideration

Not applicable

## Funding

For this research, authors did not receive any grant from any agency.

## Conflict of interest

Authors declare no conflict of interest.

## Author’s contribution

AEEH: conceptualization, design, manuscript writing, software. MN: design, data curation, manuscript writing. NUE: design, data analysis, final drafting. IHT: data curation, data analysis. MAAM: software, data analysis. SA: monitoring, draft revision. MSI: supervison, final drafting.

## Consent to publication

All authors have read the final version of the manuscript and approved the manuscript.

## Notes

### Competing Interest Statement

The authors have declared no competing interest.

